# Tobacco endgame intervention impacts on health gains and Māori:non-Māori health inequity: a simulation study of the Aotearoa-New Zealand Tobacco Action Plan

**DOI:** 10.1101/2022.07.17.22277571

**Authors:** Driss Ait Ouakrim, Tim Wilson, Andrew Waa, Raglan Maddox, Hassan Andrabi, Shiva Raj Mishra, Jennifer Summers, Coral Gartner, Raymond Lovett, Richard Edwards, Nick Wilson, Tony Blakely

## Abstract

**Objective:** To estimate the health gains and Māori:non-Māori health inequality reductions of the Aotearoa/New Zealand Government’s proposed endgame strategy.

**Design:** Simulation modelling: a Markov model to estimate future yearly smoking and vaping prevalence (for business-as-usual [BAU] and intervention scenarios) linked to a proportional multistate lifetable model with 16 tobacco-related diseases to estimate future morbidity and mortality rates, and health adjusted life years (HALYs).

**Setting:** Aotearoa.

**Participants:** Population alive in 2020 (5.08 million) simulated over the rest of their lifespan.

**Interventions:** 1. Denicotinisation of all retail tobacco in 2023, 2. 1 plus media promotion, 3. 95% reduction in tobacco retail outlets in 2023, 4. a tobacco free-generation whereby people born in 2006 and later are never legally able to purchase tobacco, 5. combined package of 2, 3 and 4.

**Main Outcome Measures:** Future smoking prevalence, deaths averted and HALYs by sex and ethnic group. Percent reduction in Māori:non-Māori all-cause mortality rate difference in 2040 under interventions compared to business as usual (BAU).

**Results:** The combined package of strategies was estimated to reduce adult smoking prevalence from 31.8% in 2022 to 7.3% in 2025 for Māori, and 11.8% to 2.7% for non-Māori. The 5% smoking prevalence target was forecast to be achieved in 2026 and 2027 for Māori males and females, respectively.

The HALY gains for the combined package (compared to BAU) over the population’s remaining lifespan was estimated to be 594,000 (95%UI: 443,000 to 738,000; 3% discount rate). The denicotinisation strategy alone achieved 97% of these HALYs, the retail strategy 19%, and tobacco-free generation 12%.

The future per capita HALY gains for the combined package for Māori were estimated to be 4.75 and 2.14 times higher than for non-Māori females and males, respectively. The absolute difference between Māori and non-Māori all-cause mortality for 45+ year olds in 2040 was estimated to be 23.4% (19.1% to 27.6%) less for females under the combined package compared to BAU, and 9.5% (7.5% to 11.3%) less for males.

**Conclusion:** A tobacco endgame strategy, especially denicotinisation, could dramatically reduce health inequities.

**Funding:** New Zealand Ministry of Health.

**Summary boxes:** *What is already known on this topic:*

- Modelling of health gains and health inequality reductions for some tobacco endgame strategies has been undertaken internationally, and specifically in Aotearoa (tobacco free generation policy, substantial reductions in the number of tobacco outlets including, a sinking lid that gradually phased out all tobacco supply between 2011 to 2025, restricting tobacco sales to pharmacies only with brief cessation advice provided to consumers). All modelling suggested that these interventions improved equity, of varying magnitude, in either smoking prevalence or health gain for Māori compared to non-Māori.
- Endgame modelling of denicotinisation has not been undertaken, alone or in combination with other interventions. The interplay of tobacco smoking and vaping has not been explicitly included in endgame modelling. The package of endgame strategies in the Aotearoa-New Zealand Government’s Smokefree Action Plan (Dec 2021) have not been modelled.

*What this study adds:*

- The Government’s package (denicotinisation of retail tobacco, 95% reduction in the number of tobacco retail outlets; and a tobacco-free generation) if implemented in 2023 is forecast to achieve less than 5% smoking prevalence by 2025 for non-Māori, and by 2027 for Māori.
- Denicotinisation is estimated to achieve the majority of the health gains.
- A 95% retail outlet reduction and a tobacco-free generation, on their own, are unlikely to achieve a 5% smoking prevalence for any sex by ethnic groups until at least 2040.
- The combined package, compared to BAU, is estimated to reduce the Māori:non-Māori gap in 45+ year old all-cause mortality in 2040 by 22.9% (95% uncertainty interval 19.9% to 26.2%) for females and 9.6% (8.4% to 11.0%) males.

## Introduction

Despite unequivocal evidence about the harm caused by commercial tobacco, it continues to be a leading cause of avoidable morbidity and mortality.^1^ Smoking prevalence in high income countries with colonial histories has steadily decreased over recent decades, but prevalence among Indigenous peoples is often substantially higher^2^ than in the general population and is a significant contributor to health inequities.^3^

Indigenous people’s experiences of colonisation include imposition of alien societal institutions, appropriation of economic resources and exposure to racism. Referred to as ‘basic causes’^4 5^, these affect access to social determinants of health (e.g., income, housing) and, via health behaviours such as smoking rates, ultimately leading to racialised health inequities. In many instances this has been compounded by the use of tobacco as a trade commodity. Since the late 19^th^ century, tobacco companies have actively exploited and promote commercial tobacco to Indigenous peoples.^2 6 7^

In 2020-21, 22.3% of Māori (the Indigenous peoples of Aotearoa/New Zealand [A/NZ]) 15 years and older smoked at least daily. This was 2.7 times the prevalence among European/Others (8.3%; NZ Health Survey).^8^

Similar to other high-income countries, A/NZ’s tobacco control programme includes: restricting the promotion of tobacco products, providing cessation support, implementing mass media campaigns, regular increases in tobacco excise tax, and smokefree area policies.^9^ Many of these measures have some reliance on individual capacity and access to resources needed to carry out desired behaviours such as quitting cigarettes. These resources are inequitably distributed across the A/NZ population, likely explaining a failure of A/NZs tobacco control programme to address smoking disparities. Concern about the slow progress in reducing smoking prevalence among Māori, led Māori political and tobacco control leaders to propose a tobacco endgame in the mid-2000s. Instead of focusing on people who smoke, they argued that the tobacco industry and the products they sell should be targeted. In 2011 the A/NZ Government committed to achieving a Smokefree country by 2025^10^ (commonly interpreted as less than 5% smoking prevalence among both Māori and non-Māori). Achieving this goal required a radical departure from business as usual (BAU) approaches ^11^, but actual tobacco control policy remained relatively unchanged. The 2010s coincided with the proliferation of alternative nicotine delivery devices and, in particular, electronic cigarettes, and introduced a discourse about ‘harm minimisation’ to the tobacco endgame debate.^12^ A more holistic notion of ‘harm’ expressed among many Māori includes addiction as well as health harm, meaning that achieving an end to both nicotine addiction as well as tobacco smoking is the desired endgame.

The A/NZ Government launched an Action Plan in late 2021 to achieve the country’s endgame objective.^13^ This plan focused on smoked tobacco and sought to bring about rapid and profound reductions in smoking prevalence, and to do so equitably such that all population groups (in particular Māori) achieve minimal smoking prevalence by 2025. Three key (‘endgame’) strategies were identified in the Action Plan to achieve this goal: denicotinising retail tobacco to non-addictive levels (e.g., ≤0.4mg nicotine/cigarette)^14^ markedly reducing retail access to tobacco, and creating a ‘Tobacco-free Generation’. The latter would be achieved by progressively raising the legal age at which tobacco can be sold to young people. These measures do not directly address basic causes or social determinants of smoking related inequities. However, they substantively circumvent the role of agency (e.g., individual access to necessary social or economic resources) in being able to quit smoking or resisting initiation. As such, they have strong potential to bring equitable change in smoking behaviour.^15^ A challenge of these types of measures is that they would act against Indigenous aspirations of empowerment and self-determination^16^ if they were enacted by a predominantly non-Indigenous government ‘on’ Māori. The Action Plan has sought to address this issue by seeking Māori engagement throughout the planning and policy development stages, including establishing a Māori Governance group.

Internationally, there is growing interest in tobacco endgame goals and strategies, with an increasing number of countries adopting endgame goals and a range of bold endgame interventions proposed. Scotland, for example, has included a strong focus on equity within their endgame goals and strategies,^17^ but, other than in A/NZ, a focus on Indigenous health inequities has not been a key objective of endgame strategies. To date, the implementation of endgame interventions has been minimal and, consequently, the evidence base of their potential effects is weak.^18^ For example, none of the endgame interventions included in the A/NZ Action Plan have been implemented at country-level, with the possible exception of substantial reductions in retail supply in Hungary.

This paper aimed to estimate the future tobacco smoking prevalence, mortality and health adjusted life year (HALY) impacts (including changes in Māori/non-Māori inequalities) of tobacco endgame strategies outlined in the A/NZ Government’s proposed Action Plan.

Specific research questions were:

1. Which endgame strategies have the potential to reduce smoking prevalence to less than 5% for all sex and ethnic groups by 2025?
2. Which endgame strategies maximally reduce Māori/non-Māori health inequalities?

We used simulation modelling to calculate these estimates, using a hierarchy of data inputs ranging from trial evidence (e.g., for very low nicotine cigarettes) to observational evidence (e.g., smokers response to what they would do in the face of policies proposed) to expert knowledge elicitation when required. Forecasting the future is uncertain. Accordingly, all input parameters have uncertainty related to them that the reader can inspect, all outputs incorporate uncertainty due to the propagated input parameter uncertainty in Monte Carlo simulation, and we tease apart which input parameter uncertainty drives most of the output uncertainty. A key principle of this study is that even if as a research community we do not have ideal data, decision makers and society need the best estimates we can produce, with appropriate depiction and caveats about inevitable uncertainty.

## Methods

We used an existing tobacco simulation model^19-21^ (rated as best of 25 tobacco models globally^22^), and expanded its capabilities to create a new model called Scalable Health Intervention Evaluation (SHINE) tobacco, which includes a Markov smoking and vaping life history model and functionality for outputting packages of interventions and mortality rates by time.

### Smoking and vaping life history model

We developed a Markov model to simulate population smoking and vaping behaviours, based on seven states (Supplementary Figure S1, Table S1): never smoker (NS), current smoker (CS), never smoker current vaper (NSCV), dual user (DU), former smoker current vaper (FSCV), former smoker and/or former vaper (FSFV), never smoker former vaper (NSFV). Movement between states are determined by transition probabilities, which reflect BAU and the potential effects of interventions (below). Initiation of smoking (transition from NS to CS or DU) and vaping (transition from NS to NSCV) was assumed to occur at age 20 years. From the age of 20 onwards, any quitting of smoking was assumed permanent, parameterised as a ‘net’ cessation rate from CS and DU to either FSCV or FSFV. For proportions of the cohort in the FSCV state, there was an annual net transition probability to FSFV, but no return flow from FSFV to FSCV. The FSFV, FSCV and NSFV states were additionally modelled as 20-year tunnel states that the cohort progressed through each year, allowing the model to identify how many years each cohort was from quitting so as to incorporate decaying impacts of smoking on disease incidence by time since quitting (see below).

To specify the transition probabilities under BAU, we first estimated future daily smoking (and vaping) rates by extrapolating trends in the 2013-14 to 2019-20 New Zealand Health Survey (NZHS) data, using a two-step regression approach: 1) a best fit regression model to historic data; and 2) a regression model on the former predictions by sex, age and ethnicity to generate annual net cessation rates by cohort as they age, and annual trends in initiation.

We then calculated annual transition probabilities to achieve these projections, starting with transition probabilities between the seven states from the United Kingdom as reported by Doan et al.^23^ (Supplementary Table S2), modifying them as required (with mathematical optimisation using Excel Solver) to meet the above projections. We performed this operation by sex and ethnicity for three age cohorts (20-24, 40-44, and 60-64), and interpolated other age cohorts.

### Proportional multistate lifetable model

A proportional multistate life table (PMSLT) was used to estimate health impacts of smoking and vaping under BAU and intervention scenarios (key input parameters in Table 1). A detailed description of the model has been provided elsewhere.^24^ Briefly, the PMSLT is composed of a main cohort lifetable, which simulates the entire A/NZ population alive in 2020 until death using projected all-cause mortality and morbidity rates by sex, age, and ethnicity (Ma ori, non-Ma ori). Thus, for the youngest people we are estimating HALYs as far out as calendar years 2131; however, we focus on the next 20 years in much of the results. In parallel, proportions of the cohort also reside in 16 subsidiary tobacco-related disease lifetables according to prevalence at baseline (i.e., start of model), and in future years based on BAU disease-specific incidence, case fatality and remission rates. Within each disease lifetable, morbidity estimates (i.e. disability rates from the NZ Burden of Disease Study^25^) are attached to prevalent cases. The tobacco-related diseases included in the model are: coronary heart disease, stroke, chronic obstructive pulmonary disease (COPD), lower respiratory tract infection (LRTI), and twelve cancers (lung, oesophageal, stomach, liver, head and neck, pancreas, cervical, bladder, kidney, endometrial, melanoma, and thyroid).

**Table 1:** Baseline and business-as-usual parameters.

| Parameter | Data Source | Trend, Uncertainty, and Scenario Analyses |
| --- | --- | --- |
| **Demography** |  |  |
| Population | Statistics New Zealand (SNZ) population estimates for 2018 by sex, age-group, and ethnicity. | Uncertainty: nil |
| **BAU – epidemiological parameters** |  |  |
| All-cause mortality rates (ACMR) | SNZ mortality rates by sex, age, and ethnicity for 2020 | Trends in ACMR were estimated using GHDx data (IMME). The annual percentage change in the age-standardised all-cause mortality rates from 1990 to 2019 was -1.9% for sexes combined. Retaining the original BODE3 model assumption of a 0.5 percent point greater APC for Māori (due to long run trends of closing ethnic inequalities in mortality), we arrived at APC's for ACMR from 2020 to 2035 of: Māori = -2.0%; non-Māori = -1.5%. They were uniform by age. No trends applied beyond 2035.<br>Uncertainty: nil. |
| All-cause morbidity rates | NZ Burden of Disease Study <sup>25</sup> | Data on years of life lived with disability (YLD) were obtained from the NZBDS for each sex and age group in 2016 and divided by the population in each sex by age by ethnic group to generate morbidity rates. No time trend was allowed. |
| Disease specific incidence, prevalence, and case-fatality rates | NZ Burden of Disease Study <sup>25</sup> | For each tobacco-related disease, coherent sets (by sex, age, and ethnicity) of incidence rates, prevalence, case-fatality rates (CFR), and remission rates (zero for non-cancers, the complement of the CFR for cancers to give the expected 5-y relative survival) were estimated using DISMOD II.<br>Cancer incidence and CFR annual percentage change (APC) trends using Poisson regression historic trends of incidence and case-fatality rates of diseases. The APCs included as inputs to the PMSLT model out to year 2035 and held constant beyond (future prevalence changes dynamically with model). It was assumed that the APCs were constant by ethnicity.<br>Uncertainty: starting in 2020, rates all +/- 5% standard deviation (SD), correlations 1.0 between four sex by ethnic group categories for all diseases. APC all +/- 0.5% SD normal, correlations 1.0 between four sex by ethnic groups for all diseases. |
| Disease specific morbidity | NZ Burden of Disease Study <sup>25</sup> | The sex and age specific disability rates were calculated as disease's YLD obtained divided by the prevalent cases.<br>The same disability rate was assumed by ethnicity (i.e., those with disease are assumed |

Health and Inequality Impacts of Tobacco Endgame Strategies in Aotearoa
| Parameter | Data Source | Trend, Uncertainty, and Scenario Analyses |
| --- | --- | --- |
|  |  | to have same severity distribution across ethnicity).<br>Uncertainty: +/- 5% SD (beta distribution) |
| **Tobacco smoking and vaping** |  |  |
| Smoking (daily) | NZ Health Survey | Logistic regression of NZ Health Survey data for years 2011 to 2019 was undertaken to ‘predict’ the prevalence of daily smoking (at least one cigarette per day) for years 2020 to 2040. This ‘prediction file’ was then re-analysed from a sex by ethnicity by five-year age group perspective (i.e., 72 separate sex by age by ethnicity cohorts) to generate future BAU smoking prevalence – and a yearly (cohort aging) rate of decline – that was then used in the exposure model. |
| Vaping (daily e-cigarette use) | NZ Health Survey | Same as above for smoking, but for ‘vaping’ at least daily. |
| **Association of smoking and vaping with disease incidence rates** |  |  |
| Smoking-disease incidence rate ratios | Relative risks of disease incidence for the association of current (or ex-smoker) with never smoker were sourced from NZ linked census-cancer<sup>33</sup> and census-mortality<sup>34</sup> (censuses include smoking question) and CPS II data for respiratory diseases.<sup>35</sup> Attenuation over time since quitting for ex-smokers was modelled using equations and coefficients from Hoogenveen et al.<sup>36</sup> | Standard errors of regression coefficients as described in Appendix C |

Within each disease lifetable, an intervention is run in parallel to BAU with different disease incidence rates given changes in smoking and vaping life histories (see next section). Each disease lifetable estimates the difference between intervention and BAU in disease mortality and morbidity rates that are then added to matching entities in the main lifetable.

### Connecting the smoking-vaping life history model to the PMSLT – using population impact fractions

For each sex by age by ethnic group, and each annual time step into the future, a population impact fraction (PIF) is calculated for each tobacco-related disease. The generic formula^26^ is:

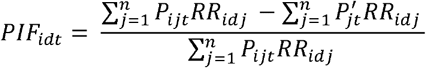

where: *i* subscripts each sex by age by ethnic group, d subscripts each disease, t subscripts each time step or yearly cycle, *j* subscripts *n* states in the smoking-vaping life history model, *RR* is the incidence rate ratio for disease d and smoking-vaping state *j*, and possible varying by demographics (e.g., by sex and age, but not by ethnic group (note the RR does not vary by time step *t*), and *P* (*P*^*’*^) is the proportion of the demographic cohort (*i*) in each of *j* states in each time step *t*. These PIFs are the percentage change in incidence rates for each smoking related disease inputted to the PMSLT.

The source and values of the tobacco-related disease incidence rates and rate ratios are given in Appendices A-B and Supplementary Tables S3-S19. Harm from vaping was modelled as 5% to 20% of tobacco harm following Mendez and Warner^27^ (Beta distribution with median 11% and 95% uncertainty interval [UI] of 5% to 20%).

### Interventions

To parameterise the intervention scenarios we adapted initial estimates by Wilson et al,^28^ which derived their potential effects based on A/NZ-specific literature (including a randomised trial of denicotinised cigarettes) and international literature; adaptation for this paper included incorporating additional research and expert judgements by the authors. The intervention specifications are shown in Table 2; below we give key parameters (and there uncertainty intervals that are sampled from in Monte Carlo simulation). Briefly:

**Table 2:** Intervention input parameter table.

| Parameter | Description |
| --- | --- |
| ***Denicotinisation*** |  |
| NS→ CS (age 20 only)<br>NS→ DU (age 20 only) | 10% (SD 5%) of BAU initiation at age 20 by five years after implementation ($X = \text{Beta}(3.6, 32.4)$, median 9.3%, 95% UI 2.6% to 21.5%). Implemented as $X^{(t/5)}$ scalar applied to the BAU initiation rates in years $t$ (1 to 5) after introduction of the policy, then held at $X\%$ thereafter. |
| CS→ FSFV<br>CS→ FSCV<br>DU→ FSFV<br>DU→ FSCV | Using an expert knowledge elicitation (see Appendix D), smoking prevalence (i.e. $X = \text{prevalence}$ in states CS and DU) with mean 15.2% (SD 7.84%, $X = \text{Beta}(3.19, 17.78)$, median 14.1%, 95% UI: 3.7% to 32.9%) of BAU smoking prevalence five years after low nicotine policy implementation, due to quitting or switching to vaping (i.e. disregarding initiation impacts that will additionally impact prevalence among 20-24 year olds in first five years of the model). Implementation was as $X^{(t/5)}$ scalar applied to BAU CS and DU prevalence where $t$ is the 1 to 5 years after intervention. For the sixth and subsequent years, the transition probabilities were twice those in BAU (due to an ongoing higher NCR, given non-addictive levels of nicotine in tobacco). |
| NS→ NSCV | No change. |
| ***Denicotinisation plus Mass media*** |  |
| NS→ CS (age 20 only) | As above for low nicotine. |
| NS→ DU (age 20 only) | As above for low nicotine. |
| NS→ NSCV (age 20 only) | No change. |
| CS→ FSFV<br>CS→ FSCV<br>DU→ FSFV<br>DU→ FSCV | As above for low nicotine from year 1 to 5 + twice the absolute contribution of the routine media/Quitline campaign to background net cessation (i.e. $1.055\% \times 2 = 2.1\%$)<sup>31</sup>. Subsequent years: transition to quitting or vaping were twice those in BAU |
| ***Retail outlet restriction to about 300 outlets (about 5% of current outlets; assumed supply of e-cigarettes reduces commensurately)***<sup>†</sup> |  |
| NS→ CS | As per the increase in cessation probabilities (CS→FSFV, etc, below), we reduced the initiation rate by $X = \text{Beta}(23.4, 97.2)$, median 19.2%, 95% UI: 12.9% to 26.9%. Applies in 2023 onwards (as youth contemplating initiating in the future confront lesser retail availability as well). |

Health and Inequality Impacts of Tobacco Endgame Strategies in Aotearoa
| Parameter | Description |
| --- | --- |
| NS→ DU | As above for NS→CS. |
| CS→ FSFV<br>CS→ FSCV<br>DU→ FSFV<br>DU→ FSCV | As a low estimate of one-off quitting, we used that from studies modelling reducing retail outlets in terms of increased travel costs <sup>37</sup>: a reduction in the prevalence of 15.6% for Māori, and 16.0% for non-Māori – or 15.8% overall.<br>As a high estimate, we used that from the NZ ITC study where – in response to a question whether they would quit in response to a 95% reduction in retail outlets - 23.0% said they would quit (half quitting → FSFV, half switching to FSCV).<sup>38</sup><br>Placing the mean at 19.4% (average of above 15.8% and 23%) and using 15.8% and 23% as one SD either side of the mean (SD = 3.6%), we parameterised the one-off increase in smoking net cessation as $X = \text{Beta}(23.4, 97.2)$, median 19.2% (i.e., percentage point increase), 95%UI: 12.9% to 26.9%. Note this increase was on top of BAU transition probabilities, and halved over CS→FSFV and CS→FSCV, and halved over DU→FSFV and DU→FSCV. E.g. if the CS→FSFV was 5%, the intervention CS→FSFV transition probability was $5\% + (1-5\%) \times 0.5 \times X\%$.<br>This effect was in the year of intervention only– in years after the retail outlet restriction, the transition probabilities out of CS and DU reverted to BAU. |
| NS→NSCV | Unchanged |
| ***Tobacco-free generation*** |  |
| Smoking initiation rate (NS→ CS; occurs only at age 20) | For two reasons, a tobacco-free generation proposal will not immediately achieve zero uptake at age 20; 1) our model for parsimony assumes all uptake at age 20, but the minimum legal age of purchasing is 18 years; 2) social supply will allow some young people to keep initiating. We therefore assumed that initiation at age 20 in our model (essentially an average of all initiation by [say] age 25) will asymptote to a mean of $X=10\%$ (SD 5%) of BAU in 10 years (Beta (3.6, 32.4), median 9.3%, 95%UI: 2.6% to 21.5%), with the scalar of BAU initiation rate of $X^{(t/10)}$ for $t = 1$ to 10 years after the tobacco-free generation policy is implemented, then $X$ of BAU initiation thereafter. |
| NS→ DU | As above for NS→CS. |
| NS→ NSCV | Unchanged<sup>†</sup> |

Health and Inequality Impacts of Tobacco Endgame Strategies in Aotearoa
| Parameter | Description |
| --- | --- |
| ***Combined: Denicotinisation + retail + tobacco-free*** |  |
| NS→ CS (age 20 only) | Cumulative impact. If the % reduction in initiation in year t for denicotinisation, retail and tobacco-free was A%, B%, and C%, then the reduction in the combined intervention was $1 - (1-A)(1-B)(1-C)$. |
| NS→ DU (age 20 only) | As above for NS→CS. |
| CS→ FSFV<br>CS→ FSCV<br>DU→ FSFV<br>DU→ FSCV | Cumulative impact. If the % increase in quitting or switching in year t for denicotinisation, media and retail was A% and B%, then the increase in the combined intervention was $1 - (1-A)(1-B)(1-C)$. |
| NS→ NSCV | Unchanged. |

NS=never smoker; CS=current smoker (but not a dual user); DU=dual user; NSCV=never smoker, current vaper; FSFV=former smoker and/or former vaper; FSCV=former smoker current vaper.
†If the availability of alternative nicotine delivery systems (ENDS, e.g. e-cigarettes) does not reduce commensurately with these policy interventions, one would expect larger switched to ANDS which would reduce smoking prevalence further (but increase DU, FSCV and possibly NSCV state prevalence). We do not model this explicitly but consider it in the Discussion.

- Denicotinisation: initiation was estimated to reduce to 10% (95% UI 2.6% to 21.5%) of that in BAU by five years after implementation; cessation transition probabilities were increased so that over five years the smoking prevalence in CS and DU states was 15.2% (95% UI 3.7% to 32.9%) of that in BAU, and from the sixth-year onward cessation transition probabilities were doubled.
- Denicotinisation plus mass media: as above, plus an extra increase in cessation rates in the first five years of 2.1% (equivalent to twice the impact of past Quitline media campaigns in A/NZ on net cessation rates).
- Retail outlet reduction: we used the average of two inputs: a) previous modelling by ourselves ^29^ of increasing travel time, converted to cost and then through price elasticities that estimates a 15.8% reduction in smoking prevalence in the year of implementation, b) 23.0% of respondents (smokers) to the NZ International Tobacco Collaboration study saying they would quit if outlets reduced by 95%. We used the average of these two (19.4%; 95% UI 12.9% to 26.9%) as the one-off increase in net cessation in the year of the policy implementation. The same magnitude reduction in initiation was included in the year of implementation and all subsequent years.
- Tobacco-free generation: in theory, initiation will reduce to zero. In practice, social supply is likely. The exact reduction in initiation is uncertain, so we specified that future initiation rates will be 10% of BAU with wide uncertainty (95%UI: 2.6% to 21.5%), achieved 10 years after the policy is introduced.

### Analyses and parameter uncertainty

We produced the following outputs. First, deaths averted by time period. Second, health-adjusted life years (HALYs; 3% annual discount rate) gained from each intervention, both the total number and age-standardised (using Māori population 2020) per 1000 people. Third, we calculated the age-standardised all-cause mortality rate differences between Māori and non-Māori (by sex) for 45+ year old (by age in the future), under BAU and each intervention, and presented the percentage difference in the rate difference for each intervention compared to BAU.

The BAU and each intervention scenario were simulated 2000 times using Monte Carlo simulation, drawing from the probability density functions specified in Tables 1 and 2.

To help understand the uncertainty in our modelling, we used univariate sensitivity analyses to depict which input parameter uncertainty generates the most uncertainty in lifetime HALY gains for all sex and ethnic groups combined for the combination endgame policy package compared to BAU. The result is presented as a ‘Tornado Plot’ showing the changes in model outputs for selecting the 2.5^th^ and 97.5^th^ percentile of each input parameter in turn (holding all other inputs at their expected value).

## Results

### Achieving < 5% prevalence

The modelled combined package achieves a profound and rapid reduction in smoking prevalence (Figure 1 and Supplementary Table S23). In 2022, the year before policy implementation, Maori age 20+ smoking prevalence is 31.8%, falling to 28.7% (95% UI: 25.5% to 30.4%) under BAU by 2025 (the year targeted to have smoking prevalence less than 5% by the A/NZ Parliament). Under the combined package Māori smoking prevalence decreases to 7.3% (3.9% to 9.2%) in 2025 (females: 8.2%, 4.3% to 10.5%; males: 6.3%, 3.4% to 7.9%). For non-Māori, the smoking prevalence is 11.8% in 2022, falling to 10.8% (95% UI: 9.6% to 11.2%) in 2025 under BAU and decreasing to 2.7% (1.4% to 3.5%) under the combined package (females: 2.5%, 1.3% to 3.3%; males: 2.9%, 1.5% to 3.8%). The combined package achieves the under 5% smoking prevalence target in 2026 and 2027 for Māori males and females, respectively.

**Figure 1.**
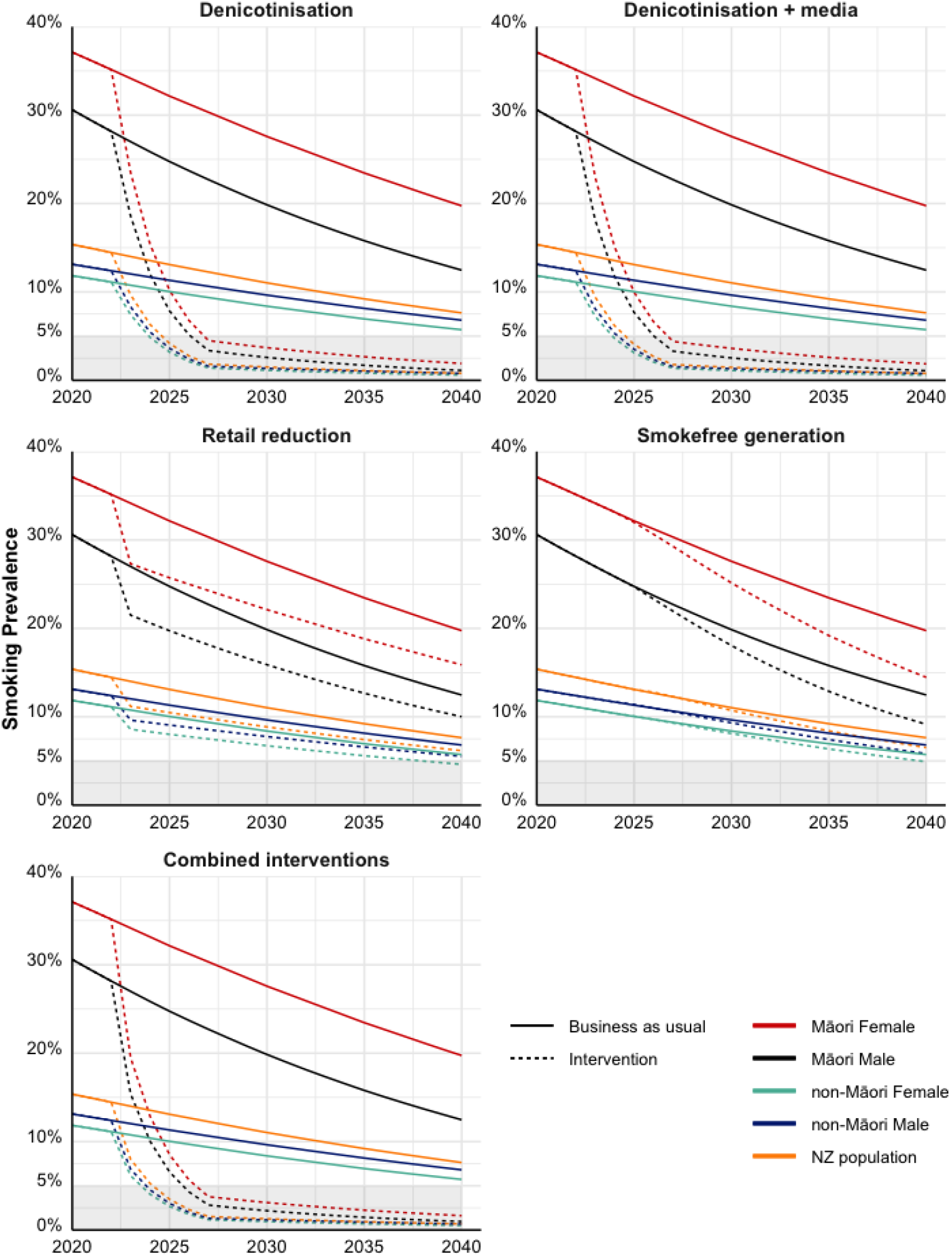
Smoking prevalence (daily, 20+ year population) in Aotearoa New Zealand under business-as-usual and interventions. Note: Prevalences are not age-standardised and are calculated for the projected age-structure of each sex by ethnic group in future years.

Denicotinisation causes the majority of forecasted decreases in smoking. Retail outlet reduction has a strong impact in its year of implementation (due to a large cessation impact), but it then tracks largely as in BAU (as no ongoing increases in cessation are assumed, and reductions in initiation take years to accrue). Neither the retail reduction nor the tobacco-free generation strategies achieve less than 5% smoking prevalence by 2025 for any sex by ethnic group.

### Deaths averted

Under the combined policy package, deaths up to 2040 were 8,150 (95%UI: 6,450 to 9,890) less than under BAU, with 27% to 30% of these averted deaths among each of female Māori, female non-Māori and male non-Māori (Table 3).

**Table 3:**
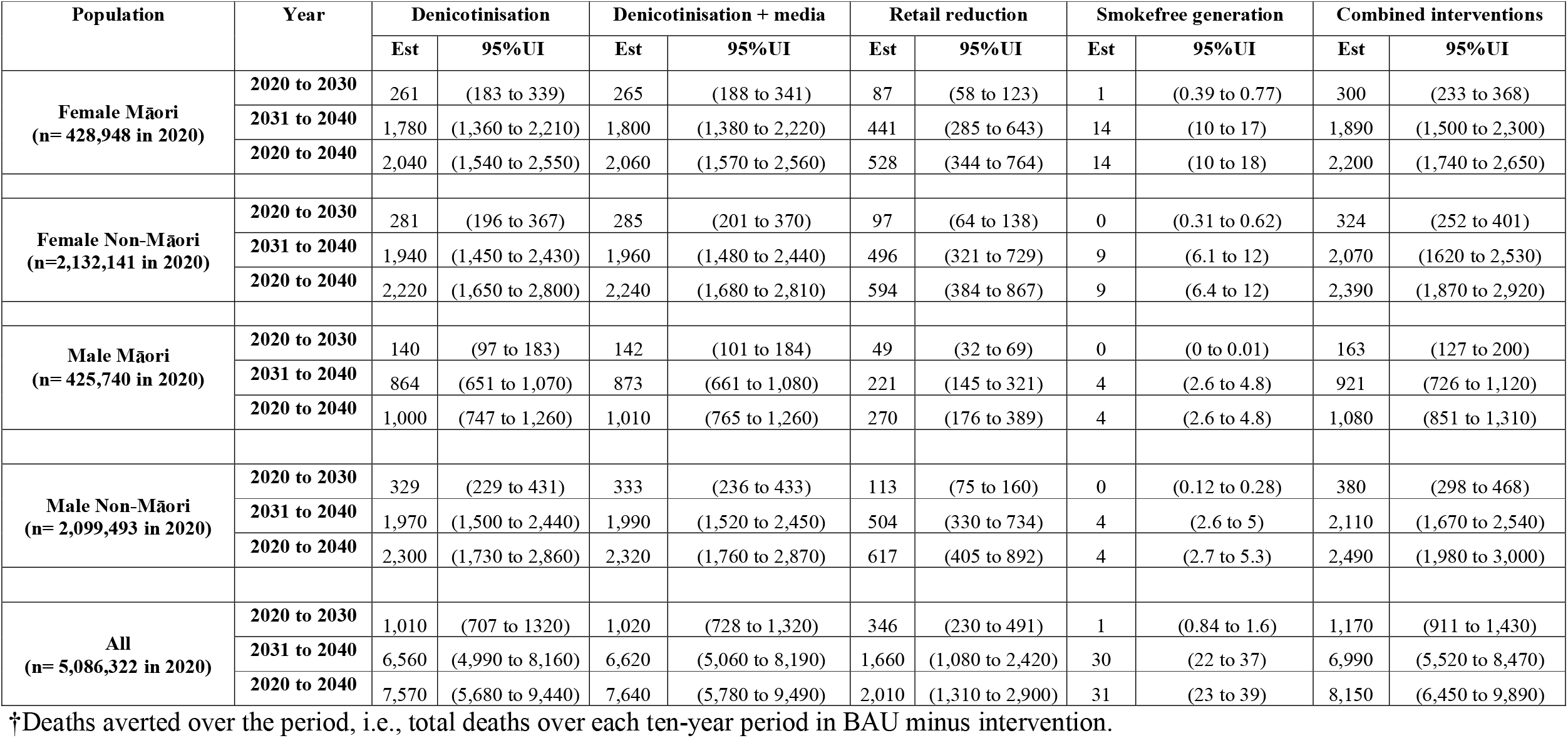
Deaths averted† during 2020-30 and 2031-40, in Aotearoa New Zealand by strategy.

| Population | Year | Denicotinisation |  | Denicotinisation + media |  | Retail reduction |  | Smokefree generation |  | Combined interventions |  |
| --- | --- | --- | --- | --- | --- | --- | --- | --- | --- | --- | --- |
|  |  | Est | 95%UI | Est | 95%UI | Est | 95%UI | Est | 95%UI | Est | 95%UI |
| **Female Māori**<br>(n= 428,948 in 2020) | **2020 to 2030** | 261 | (183 to 339) | 265 | (188 to 341) | 87 | (58 to 123) | 1 | (0.39 to 0.77) | 300 | (233 to 368) |
|  | **2031 to 2040** | 1,780 | (1,360 to 2,210) | 1,800 | (1,380 to 2,220) | 441 | (285 to 643) | 14 | (10 to 17) | 1,890 | (1,500 to 2,300) |
|  | **2020 to 2040** | 2,040 | (1,540 to 2,550) | 2,060 | (1,570 to 2,560) | 528 | (344 to 764) | 14 | (10 to 18) | 2,200 | (1,740 to 2,650) |
| **Female Non-Māori**<br>(n=2,132,141 in 2020) | **2020 to 2030** | 281 | (196 to 367) | 285 | (201 to 370) | 97 | (64 to 138) | 0 | (0.31 to 0.62) | 324 | (252 to 401) |
|  | **2031 to 2040** | 1,940 | (1,450 to 2,430) | 1,960 | (1,480 to 2,440) | 496 | (321 to 729) | 9 | (6.1 to 12) | 2,070 | (1,620 to 2,530) |
|  | **2020 to 2040** | 2,220 | (1,650 to 2,800) | 2,240 | (1,680 to 2,810) | 594 | (384 to 867) | 9 | (6.4 to 12) | 2,390 | (1,870 to 2,920) |
| **Male Māori**<br>(n= 425,740 in 2020) | **2020 to 2030** | 140 | (97 to 183) | 142 | (101 to 184) | 49 | (32 to 69) | 0 | (0 to 0.01) | 163 | (127 to 200) |
|  | **2031 to 2040** | 864 | (651 to 1,070) | 873 | (661 to 1,080) | 221 | (145 to 321) | 4 | (2.6 to 4.8) | 921 | (726 to 1,120) |
|  | **2020 to 2040** | 1,000 | (747 to 1,260) | 1,010 | (765 to 1,260) | 270 | (176 to 389) | 4 | (2.6 to 4.8) | 1,080 | (851 to 1,310) |
| **Male Non-Māori**<br>(n= 2,099,493 in 2020) | **2020 to 2030** | 329 | (229 to 431) | 333 | (236 to 433) | 113 | (75 to 160) | 0 | (0.12 to 0.28) | 380 | (298 to 468) |
|  | **2031 to 2040** | 1,970 | (1,500 to 2,440) | 1,990 | (1,520 to 2,450) | 504 | (330 to 734) | 4 | (2.6 to 5) | 2,110 | (1,670 to 2,540) |
|  | **2020 to 2040** | 2,300 | (1,730 to 2,860) | 2,320 | (1,760 to 2,870) | 617 | (405 to 892) | 4 | (2.7 to 5.3) | 2,490 | (1,980 to 3,000) |
| **All**<br>(n= 5,086,322 in 2020) | **2020 to 2030** | 1,010 | (707 to 1320) | 1,020 | (728 to 1,320) | 346 | (230 to 491) | 1 | (0.84 to 1.6) | 1,170 | (911 to 1,430) |
|  | **2031 to 2040** | 6,560 | (4,990 to 8,160) | 6,620 | (5,060 to 8,190) | 1,660 | (1,080 to 2,420) | 30 | (22 to 37) | 6,990 | (5,520 to 8,470) |
|  | **2020 to 2040** | 7,570 | (5,680 to 9,440) | 7,640 | (5,780 to 9,490) | 2,010 | (1,310 to 2,900) | 31 | (23 to 39) | 8,150 | (6,450 to 9,890) |

†Deaths averted over the period, i.e., total deaths over each ten-year period in BAU minus intervention.

### HALYs gained

For the combined intervention compared to BAU, by sex and ethnic group, 28% to 30% of all HALYs gained by the combined package were among female Māori, female non-Māori and male non-Māori, with a lesser 14% among male Māori. For sexes and ethnic groups combined, and for the remainder of the lifespan of the population alive in 2020, there was an estimated 594,000 HALYs gained (95% UI: 443,000 to 738,000: bottom right of Table 4). The majority (90%) of these HALYs gained were after 2040.

**Table 4:** Health gain (in HALYs gained) for people alive in 2020 (base-year, N = 5,086,322) in Aotearoa New Zealand by the model led policies, by timeline into the future (3% discount rate)

| Population | Year | Denicotinisation |  | Denicotinisation + media |  | Retail reduction |  | Smokefree generation |  | Combined interventions |  |
| --- | --- | --- | --- | --- | --- | --- | --- | --- | --- | --- | --- |
|  |  | Estimate | 95% UI | Estimate | 95% UI | Estimate | 95% UI | Estimate | 95% UI | Estimate | 95% UI |
| Female Māori | 2020 to 2030 | 955 | (679 to 1,260) | 971 | (699 to 1,260) | 335 | (221 to 475) | 23 | (16 to 31) | 1,130 | (884 to 1,380) |
|  | 2031 to 2040 | 10,600 | (7,990 to 13,100) | 10,700 | (8,160 to 13,100) | 2,800 | (1,830 to 3,990) | 325 | (242 to 406) | 11,500 | (9,370 to 13,700) |
|  | 2041 to 2131 | 151,000 | (111,000 to 188,000) | 151,000 | (112,000 to 189,000) | 24,800 | (15,900 to 36,700) | 29,900 | (19,400 to 40,600) | 157,000 | (116,000 to 195,000) |
|  | All | 162,000 | (120,000 to 202,000) | 163,000 | (121,000 to 203,000) | 27,900 | (18,000 to 41,100) | 30,300 | (19,700 to 41,000) | 170,000 | (127,000 to 210,000) |
| Female Non-Māori | 2020 to 2030 | 1,450 | (1,030 to 1,940) | 1,470 | (1,050 to 1,950) | 525 | (346 to 745) | 23 | (16 to 32) | 1,710 | (1,330 to 2,130) |
|  | 2031 to 2040 | 14,700 | (11,000 to 18,300) | 14,800 | (11,200 to 18,400) | 3,990 | (2,610 to 5,770) | 334 | (239 to 443) | 16,000 | (12,800 to 19,200) |
|  | 2041 to 2131 | 142,000 | (104,000 to 182,000) | 143,000 | (105,000 to 182,000) | 28,200 | (17,800 to 42,800) | 15,300 | (9,950 to 22,400) | 149,000 | (109,000 to 189,000) |
|  | All | 159,000 | (116,000 to 201,000) | 160,000 | (117,000 to 202,000) | 32,700 | (20,800 to 49,200) | 15,600 | (10,300 to 22,900) | 166,000 | (124,000 to 209,000) |
| Male Māori | 2020 to 2030 | 596 | (423 to 786) | 606 | (436 to 792) | 214 | (141 to 303) | 17 | (12 to 22) | 707 | (554 to 871) |
|  | 2031 to 2040 | 6,080 | (4,570 to 7,520) | 6,140 | (4,670 to 7,560) | 1,640 | (1,070 to 2,350) | 230 | (171 to 291) | 6,650 | (5,360 to 7,940) |
|  | 2041 to 2131 | 70,100 | (49,400 to 90,600) | 70,500 | (49,600 to 91,000) | 12,400 | (7,990 to 18,800) | 12,800 | (8,220 to 18,100) | 73,600 | (52,100 to 94,200) |
|  | All | 76,700 | (54,800 to 98,300) | 77,200 | (55,300 to 98,800) | 14,300 | (9,240 to 21,400) | 13,000 | (8,450 to 18,300) | 80,800 | (58,200 to 103,000) |
| Male Non-Māori | 2020 to 2030 | 1,620 | (1,140 to 2,150) | 1,640 | (1,170 to 2,160) | 585 | (384 to 830) | 24 | (16 to 33) | 1,910 | (1,500 to 2,360) |
|  | 2031 to 2040 | 15,900 | (12,000 to 19,700) | 16,000 | (12,200 to 19,800) | 4,300 | (2,810 to 6,180) | 345 | (249 to 455) | 17,300 | (14,000 to 20,800) |
|  | 2041 to 2131 | 151,000 | (112,000 to 193,000) | 152,000 | (112,000 to 193,000) | 29,500 | (18,900 to 44,400) | 16,300 | (10,800 to 23,800) | 158,000 | (119,000 to 199,000) |
|  | All | 168,000 | (126,000 to 214,000) | 169,000 | (127,000 to 214,000) | 34,400 | (22,100 to 51,400) | 16,700 | (11,100 to 24,300) | 177,000 | (134,000 to 221,000) |
| All population | 2020 to 2030 | 4,630 | (3,260 to 6,140) | 4,690 | (3,360 to 6,170) | 1,660 | (1,090 to 2,360) | 88 | (60 to 118) | 5,460 | (4,280 to 6,740) |
|  | 2031 to 2040 | 47,400 | (35,700 to 58,500) | 47,800 | (36,400 to 58,700) | 12,700 | (8,300 to 18,300) | 1,230 | (909 to 1,590) | 51,500 | (41,500 to 61,600) |
|  | 2041 to 2131 | 514,000 | (378,000 to 650,000) | 517,000 | (380,000 to 653,000) | 94,800 | (60,900 to 143,000) | 74,200 | (49,000 to 104,000) | 537,000 | (396,000 to 673,000) |
|  | All | 566,000 | (421,000 to 711,000) | 569,000 | (424,000 to 714,000) | 109,000 | (70,200 to 163,000) | 75,500 | (50,200 to 105,000) | 594,000 | (443,000 to 738,000) |

The denicotinisation strategy alone achieves 97% of the HALYs of the combined package, retail outlet reduction alone 19% and the tobacco-free generation alone 12%. For the tobacco-free generation, the vast majority (98%) of HALYs gained over the lifespan of the population occurred after 2040. Supplementary Figure S3 provides a comparison in terms of health gains from the endgames strategies evaluated in this paper with other large scale public health policies (modelled or already in place) in A/NZ.

### Inequality impacts

Figure 2 shows the ratio of age-standardised per capita HALYs gains for Māori compared to non-Māori. For the combined package, Māori females gained 4.75 times as many HALYs per capita as non-Māori females, and Māori males gained 2.15 times as many as non-Māori males. The Māori:non-Māori ratio of per capita HALY gains was similar for other interventions, except it was higher for the tobacco-free generation (noting, though, that the absolute gains were less for this strategy – see Supplementary Table S24).

**Figure 2:**
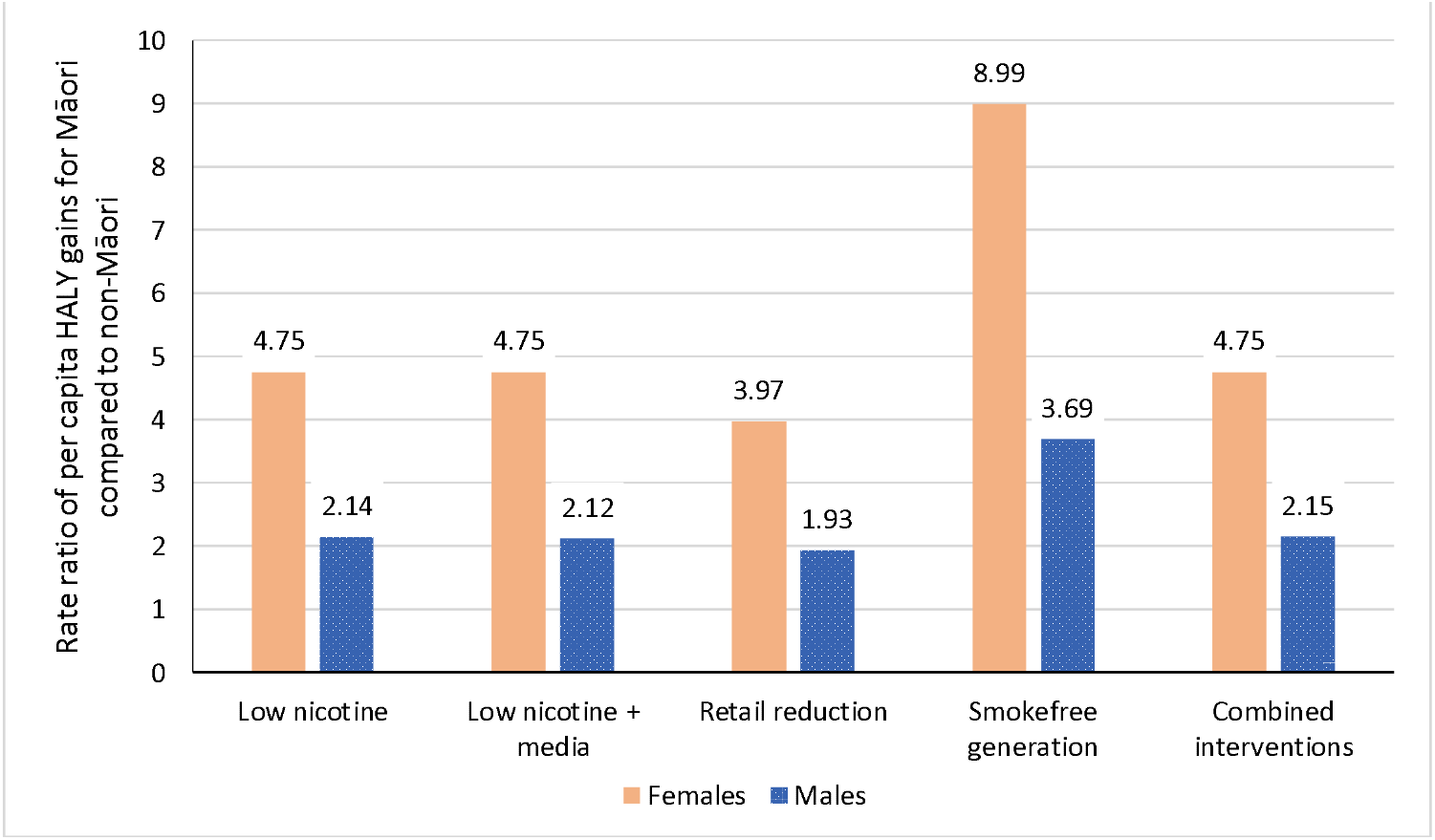
Ratios of per capita HALY gains over the remainder of the 2020 Aotearoa-New Zealand population’s lifespan, for Māori compared to non-Māori. Calculated using cohorts defined by age in 2020, age standardised using the 2020 Māori population.

Māori 45+ years mortality rates in 2040 are 11.6% and 5.2% lower under the combined package than under BAU, for females and males respective. For non-Māori, these reductions are less at 2.8% and 2.3%, for females and males respectively. The impact of the combined endgame strategies on the Māori compared to non-Māori ‘gap’ (absolute difference) in mortality rates by 2040 is shown in Figure 3. The rate difference is 23.4% (95% UI: 19.1% to 27.6%) less for females for the combined package compared to BAU, and 9.5% (95% UI: 7.5% to 11.3%) less for males. The denicotinisation policy alone achieves most of this mortality rate inequality reduction, and the retail reduction strategy about a quarter of that for the combination strategy.

**Figure 3:**
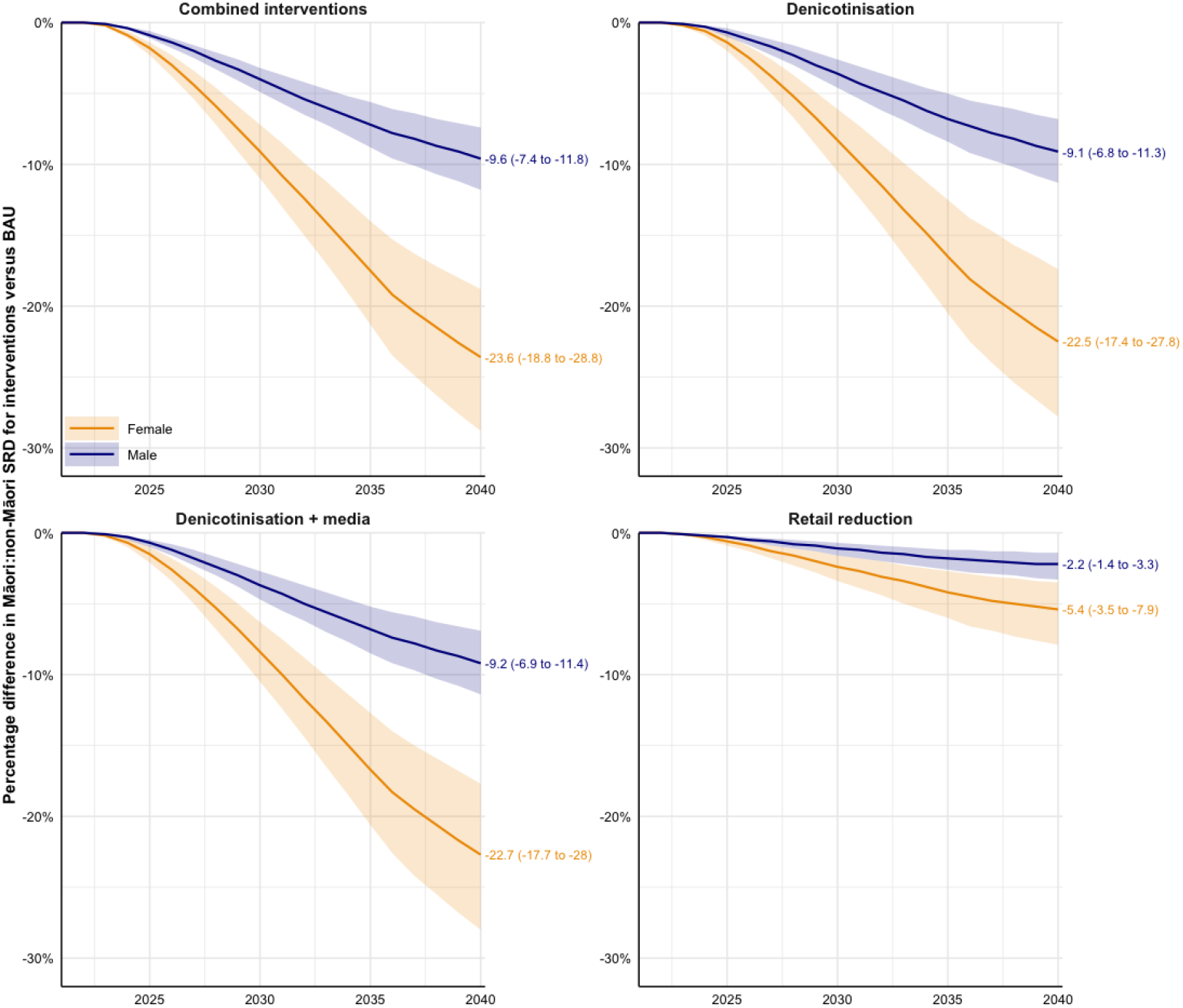
Projected percentage changes in age-standardised all-cause mortality rate differences (≥ 45 years) between Māori and non-Māori, for endgame strategies †compared to BAU. SRD: Standardised rate difference. Rates are standardised to the Māori population † We do not show the tobacco-free generation as there is no change in 45 plus year old mortality rates in this timeline.

### Sensitivity analyses

Figure 4 shows a Tornado plot of how much variation in lifetime HALYs gained (combined endgame policy; 3% discount rate) resulted from univariate sensitivity analyses about the key intervention parameters. Uncertainty about the cessation rate due to denicotinisation was clearly the major source of overall uncertainty in HALYs gained: the 97.5^th^ percentile value of increased cessation leading to 32.9% of BAU smoking prevalence (or conversely a 67.1% reduction in smoking prevalence due to increased cessation) led to 545,000 HALYs gained (end of blue bar in Figure 4) compared to 653,000 HALYs gained (end of red bar) for the 2.5^th^ percentile value of 3.7% of BAU smoking prevalence due to cessation (or conversely a large 96.3% reduction in smoking prevalence due to increased cessation). Uncertainty about other key input parameters generate considerably less uncertainty in the HALYs gained.

**Figure 4:**
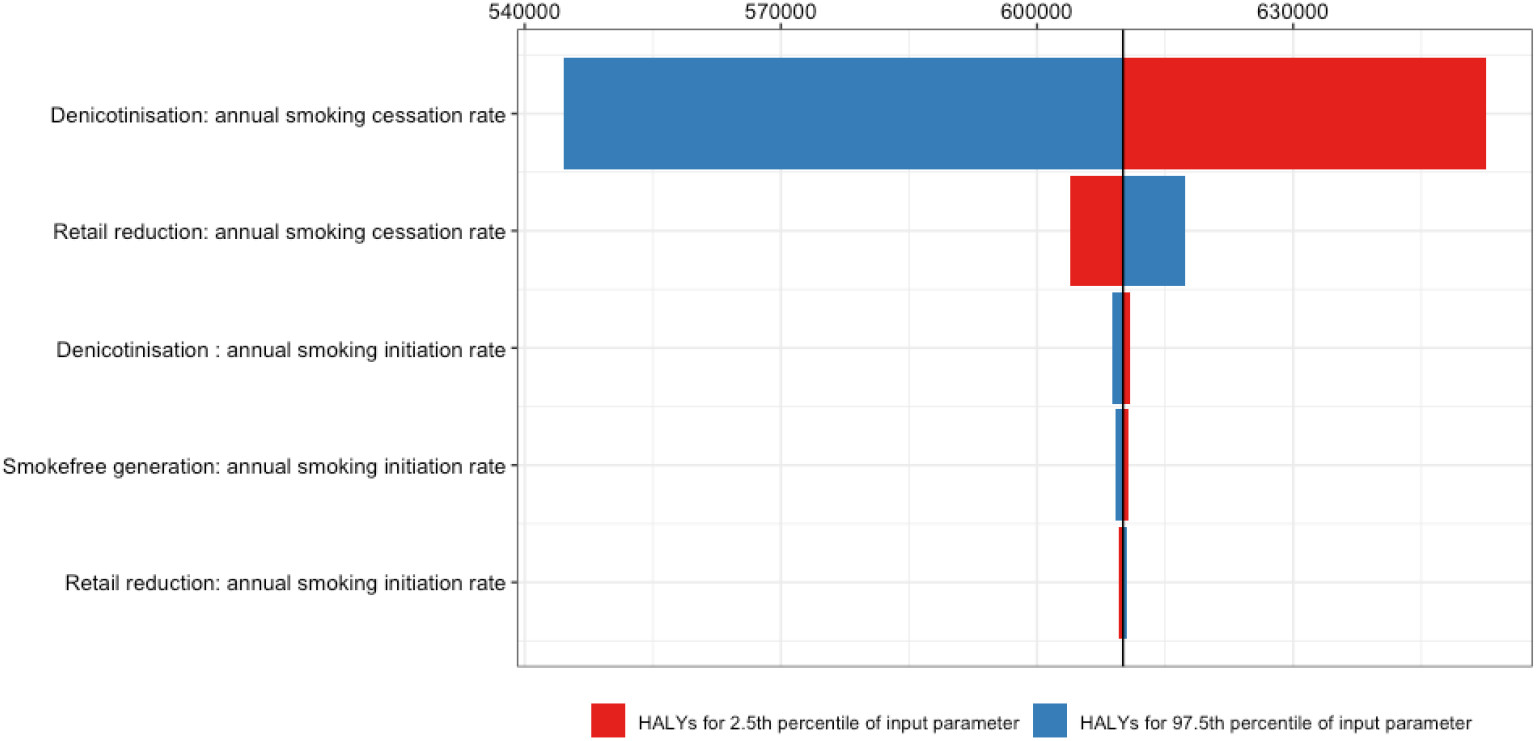
Tornado plots of total HALYs (3% discount, lifetime) showing the ranking of smoking initiation and cessation parameters by how much uncertainty their 2.5^th^ and 97.5^th^ percentiles cause for the combined intervention policy package compared to BAU, for: Percentile values for sensitivity analyses (taken from Table 2):

- *Denicotinisation: annual smoking cessation rate*. Cessation rates in first five years after policy implementation set to achieve 3.7% (2.5^th^ percentile) or 32.9% (97.5^th^ percentile) of BAU smoking prevalence [median = 14.1%].
- *Retail reduction: annual smoking cessation rate*. One off (in year of implementation) increase in cessation rate of 12.9 percentage points (2.5^th^ percentile) or 26.9 percentage points (97.5^th^ percentile) [median = 19.2 percentage points]. Note: when combined with denicotinisation (above), it acts on top of the ‘new’ (not BAU) denicotinisation cessation rate.
- *Denicotinisation: annual smoking initiation rate*. Initiation rate five years after policy implementation reduced to 2.6% (2.5^th^ percentile) or 21.5% (97.5^th^ percentile) of BAU initiation rates [median = 9.3%].
- *Smokefree generation: annual smoking initiation rate*. Initiation rate five years after policy implementation reduced to 2.6% (2.5^th^ percentile) or 21.5% (97.5^th^ percentile) of BAU initiation rates [median = 9.3%]. Note: when combined with denicotinisation (above), it acts on top of the ‘new’ (not BAU) denicotinisation cessation rate.
- *Retail reduction: annual smoking initiation rate*. Permanent decrease in initiation rate of 12.9% (2.5^th^ percentile) or 26.9% (97.5^th^ percentile) [median = 19.2%]. Note: when combined with denicotinisation and smokefree generation (above), it acts on top of the ‘new’ (not BAU) initiation rate. Note the vertical black line of 610,073 HALYs is for the median value of all input parameters. It differs modestly from the 594,000 central estimate of HALYs in the main analyses, which is the median across all iterations of the Monte Carlo analyses.

Discussion

In Aotearoa-New Zealand (A/NZ), a post-colonial country with a high smoking rates among the Indigenous Māori, we found that tobacco endgame strategies outlined in the December 2021 A/NZ Smokefree Plan ^13^, in particular denicotinisation of commercial tobacco, could have a profound positive impact on the health of Māori and notably reducing health inequity between Māori and non-Māori. For example, by 2040, a combined package including denicotinisation plus media, 95% reduction in retail outlets and a tobacco-free generation would – we estimate – reduce the gap in the mortality rate of people aged 45 years and older by 23.4% (95% UI 19.1% to 27.6%) for females and 9.5% (7.5% to 11.3%) for males, compared to ongoing BAU. It is unlikely that any other feasible health intervention would reduce ethnic inequalities in mortality by as much.

Our forecasts suggest mandating denicotinisation would have an immediate, marked and enduring impact on smoking prevalence in A/NZ. Importantly the impacts of this measure would make a significant contribution towards eliminating smoking prevalence inequities between Māori and non-Māori populations. Reducing retail access would have a lesser impact on overall prevalence and inequities and introducing a tobacco-free generation alone would take many years to take full effect with impact on smoking prevalence and then health gains. Nevertheless, the impacts of both of these measures are on par with tobacco tax increases,^30^ and greater than interventions such as mass media and quit programmes alone.^31^

The profound impact of tobacco endgame strategies on ethnic health inequalities in A/NZ shown in the model is due to higher smoking rates among Māori (especially females), but also because the smoking-related disease rates are higher among Māori (for both tobacco and non-tobacco-related reasons). Such patterning by indigeneity, ethnicity and socioeconomic position occurs in many other countries, suggesting tobacco endgame strategies will notably reduce health inequities in other countries – as well as improving the health of all citizen groups.

Tackling tobacco is not only a health issue, it has also a social and economic priority for Indigenous peoples.^32^ Whilst not presented in this paper, modelling we conducted for the A/NZ Government to underpin the Action Plan estimated income gains of US$ 1.42 billion by 2040 (3% discount rate) due to the income gains occurring among those not dying prematurely or developing chronic disease, a fillip to the A/NZ productivity and GPD overall but also a pro-equity economic boost for Māori communities.

Colonisation is an underlying driver of ethnic inequalities in smoking behaviour. Māori engagement and leadership throughout the process of developing and subsequent implementation of A/NZs Action Plan has been essential to ensure the Plan itself is not a further expression of coloniality. Legislation for the actual implementation of the Plan is expected to happen during 2022 with different measures coming into force over the next few years.

Other than a temporary ban on tobacco sales in Bhutan, no country has implemented any of the endgame interventions proposed in the A/NZ Action Plan. This lack of evidence about the real-world impacts of endgame strategies means that modelling studies’ assumptions about likely impact are based on theory, logic, expert views and simulation studies. It is therefore imperative that where endgame strategies are implemented, they are robustly evaluated to provide evidence to better inform decision-making and improve modelled estimates such as in this current study. Second, such evaluations should include a thorough investigation of equity issues, including where applicable, the exploration of intended and unintended impacts on Indigenous peoples. Thirdly, the striking equity impacts of endgame interventions estimated in this study underline that future tobacco control modelling studies should explore impacts on inequities in smoking prevalence and smoking-related disease.

### Strengths and limitations

Given data limitations, expert judgement and estimates from scenario studies were used in specifying the impacts of endgame policies. We specified substantial uncertainty about most of these inputs (Table 2), then used Monte Carlo simulations to generate uncertainty about the outputs of HALYs gained and mortality impacts. The uncertainty intervals of the HALYs, for example, are non-overlapping between the denicotinisation and retail interventions, and with BAU, suggesting a strong degree of confidence in the likely magnitude of health gains and inequality impacts. Univariate sensitivity analyses for the combined interventions policy package (Figure 4) clearly show that uncertainty about how much cessation will reduce with a denicotinisation policy is the key uncertainty in our modelling. That said, even for this cessation impact varying widely from a 67.1% to 96.3% reduction in prevalence, the lifetime HALY gains were always substantial (range 545,000 to 653,000).

We used 2013-14 to 2019-20 health survey data to parametrise our smoking-vaping life history model. Since then, 2020-21 data has found a marked drop in smoking prevalence in all socio-demographic groups. If this drop is not just a statistical anomaly, then we may have over-estimated smoking rates in the base year and BAU in the future, therefore overestimating HALY gains and mortality rate reductions arising from endgame strategies. Second, we assumed all uptake occurred at age 20, and report smoking prevalence for 20+ year olds; had we used 15+ year olds as our denominator, the smoking prevalence results reported would have been lower.

A/NZ has a fairly liberal access to alternative nicotine products, such as vaping. This meant that in our modelling, some people quitting tobacco took up vaping for a while at least (Supplementary Table S24). The generalisability of our study to other countries will depend partly on their regulatory environment for vaping.

Homegrown tobacco for personal recreational use, and illicit supply, may provide some alternative tobacco source in A/NZ with denicotinisation or substantial reduction in retail access. However, homegrown tobacco is uncommon in A/NZ due to a non-ideal physical environment in most of A/NZ for growing, and tight border security in an island nation with no land borders reduces the potential of an illicit market.

We have highlighted the importance of Māori and Indigenous engagement in the development and implementation of A/NZ’s Action Plan. The Plan also draws attention to the need for research and evaluation to provide an accountability mechanism to Māori. In this paper we attempted to uphold Indigenous Data Sovereignty principles, including Māori and other First peoples contributors (AW, RM and RL), providing data analysed against Indigenous population norms and including Indigenous interpretations. But more should be done in the future to engage Maori governance of research alongside the implementation of the Action Plan, facilitating Māori researchers undertaking that research where practicable, and prioritizing dissemination of findings to Māori communities first.

### Conclusion

Many countries have Indigenous, ethnic and socio-economic inequalities in tobacco use. This modelling study suggests that tobacco endgame strategies could have major impacts both on overall health status, and on reducing inequalities in health.

## Data Availability

All data produced in the present study are available upon reasonable request to the authors

## Statements

### Ethics

Ethics approval was not required for this study, as only secondary data sources were used.

### Transparency statement

The lead author (DAO) and senior author (TB) jointly guarantee that the manuscript is an honest, accurate, and transparent account of the study being reported. In addition, the authors indicate that no important aspects of the study have been omitted, and that any discrepancies from the study as originally planned (and, if relevant, registered) have been explained.

This research is funded by the NZ Ministry of Health and has the potential to dramatically affect the health of people in Aotearoa, especially Māori. Accordingly, additional transparency steps have taken place. These include: an extensive dialogue with the Ministry of Health staff and the NZ Associate Minister of Health on findings (they have not contributed to write-up), earlier results from this modelling have been used as a regulatory impact statement accompanying a policy proposal to Cabinet, and early dissemination with key Māori stakeholder groups in Aotearoa and international tobacco control researchers has been undertaken.

### Contributorship

Our team brings Māori lived experience (AW), Indigenous lived experience (RM, RL), and experience in research on tobacco inequalities (AW, RM, CG, RL, RE, NW, TB).

DAO, JS, NW and TB led the conceptualisation of the computer simulation modelling with data and other input specified by TW, HA and SRM. DAO and TW led the analyses and production of outputs, tables and figures. All authors contributed to data interpretation. AW, RM and RL initiated the drafting of the Introduction and Discussion, TB led the Methods and Results, and DAO led the Appendix. All authors revised the draft manuscript critically for important intellectual content.

### Role of the funding source

The views expressed in this report are solely the responsibility of the authors and they do not necessarily reflect the views, decisions, or policies of the institutions with which they are affiliated, of the NZ Ministry of Health or Government.

Specifically, the NZ Ministry of Health:

- funded the initial analyses
- did not contribute to drafting this paper
- did contribute to conceptualization of the interventions, as they are integral to the NZ Action Plan
- did provide NZ Health Survey data to parametrise the smoking-vaping life history Markov model.

## Acknowledgements

NA

## Exclusive Licence statement

I, the Submitting Author has the right to grant and does grant on behalf of all authors of the Work (as defined in the <u>author licence</u>), an exclusive licence and/or a non-exclusive licence for contributions from authors who are: i) UK Crown employees; ii) where BMJ has agreed a CC-BY licence shall apply, and/or iii) in accordance with the terms applicable for US Federal Government officers or employees acting as part of their official duties; on a worldwide, perpetual, irrevocable, royalty-free basis to BMJ Publishing Group Ltd (“BMJ”) its licensees.

The Submitting Author accepts and understands that any supply made under these terms is made by BMJ to the Submitting Author unless you are acting as an employee on behalf of your employer or a postgraduate student of an affiliated institution which is paying any applicable article publishing charge (“APC”) for Open Access articles. Where the Submitting Author wishes to make the Work available on an Open Access basis (and intends to pay the relevant APC), the terms of reuse of such Open Access shall be governed by a Creative Commons licence – details of these licences and which licence will apply to this Work are set out in our licence referred to above.

